# Fixed-Life or Rechargeable Battery for Deep Brain Stimulation: Preference and Satisfaction in Chinese Patients with Parkinson’s Disease

**DOI:** 10.1101/2020.04.28.20082677

**Authors:** Xian Qiu, Tingting Peng, Zhengyu Lin, Kaiwen Zhu, Yuhan Wang, Bomin Sun, Keyoumars Ashkan, Chencheng Zhang, Dianyou Li

**Affiliations:** Department of Neurosurgery, Ruijin Hospital, Shanghai Jiao Tong University School of Medicine, Shanghai, China; Center for Functional Neurosurgery, Ruijin Hospital, Shanghai Jiao Tong University School of Medicine, Shanghai, China; School of Nursing, Shanghai Jiao Tong University, Shanghai, China; Department of Neurosurgery, King’s College Hospital, London, UK

## Abstract

**Objective:** To evaluate the preference and satisfaction in the Chinese Parkinson’s disease (PD) patients treated with deep brain stimulation (DBS).

**Background:** DBS is a widely used therapy for PD. There is now a choice between fixed-life implantable pulse generators (IPGs) and rechargeable IPGs, each having their advantages and disadvantages.

**Methods:** Two hundred and twenty PD patients treated with DBS completed a self-designed questionnaire to assess long-term satisfaction and experience with the type of battery they had chosen, and the key factors affecting their choices. The survey was performed online and double-checked for completeness and accuracy.

**Results:** The median value of follow-up length was 18 months. The most popular way for patients to learn about DBS surgery was through media (79/220, 35.9%), including the Internet and television programs. 87.3% of the DBS used rechargeable IPGs (r-IPG). The choice between rechargeable and non-rechargeable IPGs was significantly associated with the patient’s affordability 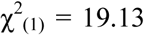, p < 0.001). Interestingly, the feature of remote programming significantly affected patients’ choices between domestic and imported brands 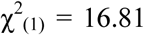, p < 0.001). 87.7% of the patients were satisfied with the stimulating effects as well as the implanted device. 40.6% of the patients with r-IPGs felt confident handling their devices within one week after discharge. More than half of the patients checked their batteries every week. The mean interval for battery recharge was 4.3 days. 57.8% of the patients spent around one-hour recharging, and 71.4% of them recharged the battery independently.

**Conclusion:** Most patients were satisfied with their choices of IPGs. The patients’ financial status and remote programming function were the two most critical factors in their decision. The skill of using rechargeable IPG was easy to master by most patients.

## Introduction

Deep brain stimulation (DBS) is a well-established neurosurgical treatment for advanced Parkinson’s disease (PD), which achieves its effects by continuous electrical current delivery to the target brain area. The power comes from an implantable pulse generator (IPG), which was initially designed to be non-rechargeable. Depending on the diagnosis and parameter settings, these fixed-life batteries had to be replaced three to five years after implantation. As the continuous stimulation calls for high energy consumption, battery depletion is the most common reason for further surgery in DBS patients. ^1,2^ Despite the battery replacement is a minor surgical procedure compared with the primary operation, several problematic issues such as infection risk and wound healing may occur after several IPG replacements. ^3^ Rechargeable batteries were introduced in 2008. Compared to fixed-life ones, rechargeable batteries offer several advantages, including smaller profile but longer battery life, requiring fewer battery replacement surgeries, and fewer battery exhausted–related problems (e.g., injuries caused by worsening of symptoms). ^4^ Research on the utility of rechargeable batteries for DBS have demonstrated adequate tolerability, a high patient satisfaction rate, and a favorable cost-effectiveness. However, patients are required to check the battery status and regularly recharge by using a handheld device. The frequency and duration of the recharge procedure depend on the power consumption of each patient. Although it is not difficult to perform, the recharge procedure can be challenging for PD patients as most of them are the elderly with various levels of cognitive deficits. ^5^

In addition to battery options, rapid technological development in the last decade has made neurologists and neurosurgeons—face the challenge of selecting an ideal individualized DBS system based on multiple variables such as lead geometry and different programming platforms.^6^ Factors including DBS target, the amount of current needed, patient’s overall health condition and their ability to deal with interactive devices and recharging, social support network, and personal financial status are of prime importance in delivering individualized therapy to patients. Another critical area that has not been fully explored is the influence of patient’s individual preferences and lifestyle concerns on the choice of the DBS device. The patient’s decision on DBS battery choice and the influencing factors of the decision have only been investigated in a few studies in Europe. ^5,7–9^

Neuromodulation with implantable devices is one of the most technologically driven disciplines within neurosurgery. In recent years, there has been a considerable expansion in choices of available DBS hardware allowing for more personalized therapies, maximizing the benefits and acceptability, whilst minimizing the risks. At the same time, DBS specialists also face the challenge of selecting among a mass of options to choose the most appropriate device for each patient. Understanding why and how patients make their choices is critical not only to improve patient acceptability and satisfaction of the therapy but also to innovate the next generation devices. In China, three DBS manufacturers (Medtronic, PINS and SceneRay) are currently offering non-rechargeable (nr-IPG) and rechargeable IPGs (r-IPG) from 1998 and 2013, 2013 and 2014, 2016 and 2019, respectively (Supplementary Figure 1). In this article, we sought to report patients’ satisfaction and preferences for the type of IPGs they chose.

## Method

### Participants

Patients diagnosed with idiopathic PD who had undergone DBS surgery in the Department of Functional Neurosurgery in Ruijin Hospital Affiliated to Shanghai Jiao Tong University School of Medicine were invited to participate in this survey. During routine clinical practice, our patients were offered a choice of r-IPG or nr-IPG, as well as of international or domestic brand, after a comprehensive and detailed explanation of each option. Patients implanted with either nr-IPG or r-IPG were included in this study. Totally 768 patients with PD who had received DBS surgeries in our center were contacted for this study.

### Questionnaire

An internet-based questionnaire (powered by www.wjx.cn) was developed and was distributed via an online chat software Wechat. The questions comprised of various aspects of the DBS devices that the patients had chosen, including (but not limited to) patients’ demographics, factors that impacted patients’ choices, patients’ satisfaction about their choices and DBS surgery. In particular, several questions were designed specifically for patients with an r-IPG device: the feasibility and reliability of the battery recharge, the interval and duration of the recharge, and the convenience of post-operative management of r-IPG. All patients provided their consent for data collection and analysis at the beginning of the questionnaire before study participation. This study was approved by Ruijin Ethical Committee.

### Statistical analysis

Data were analyzed with the SPSS software (Version 23.0. Amonk, NY: IBM Corp.). Continuous variables were expressed as Mean ± SD or median value with interquartile range (IQR). Categorical variables were presented as frequencies (%). We used Fisher’s exact test to assess the–relationship between patient’s affordability and the choice of r-IPG versus nr-IPG. We used Pearson’s or Yates’ continuity corrected Chi-square test to evaluate the influencing factors for patients’ choices of international versus domestic manufacturers in r-IPG group, and to compare the satisfaction rate between patients with r-IPG and nr-IPG. A p-value < 0.05 was considered significant.

## Results

### Demographic data

220 patients (135 men and 85 women) with PD completed the survey and were available for analysis. The response rate was 28.6% (220/768). Among them, 192 (87.3%) patients were implanted with r-IPGs and 28 (12.7%) patients with nr-IPGs. 142 (64.5%) patients had chosen a device from the international manufacturer (i.e., Medtronic) and 78 (35.5%) patients had chosen a device from domestic manufacturers (i.e., PINS or SceneRay). Overall, the mean age was 62.8 ± 9.8 years. The median value of the follow-up length was 18 (IQR: 8–36) months. Demographic data was listed in Table 1.

**Table 1.** Demographic data.

| **Characteristics** | **Total** | **r-IPG** | **nr-IPG** |
| --- | --- | --- | --- |
| **Gender** |  |  |  |
| Men | 135 | 114 | 21 |
| Women | 85 | 78 | 7 |
| **Age, year (Mean ± SD)** | 62.8 ± 9.8 | 62.5 ± 9.8 | 64.4 ± 10.0 |
| **Follow-up, month (Median, IQR)** | 18 (8–36) | 19 (9–38) | 15.5 (5.25–26.5) |
| **Manufacturer** |  |  |  |
| Medtronic | 142 | 136 | 6 |
| PINS & SceneRay | 78 | 56 | 22 |

### Source of DBS information

The web and television media and doctor’s referrals served as two important tunnels for PD patients to know DBS before seeking surgery [36.1% (79/219) and 35.6% (78/219), respectively]. The remaining quarter of the patients (25.5%, 56/219) came to our center learned the information about DBS by word of mouth from other patients (Table 2).

**Table 2.** Approaches to learn about deep brain stimulation. (N = 219)^*^

| **Approaches** | **Number (%)** |
| --- | --- |
| Media (i.e., Web and television) | 79 (36.1%) |
| Doctor's referrals | 78 (35.6 %) |
| Word of mouth from patients | 56 (25.6%) |
| Others | 6 (2.7%) |

\*One missing data.

### Influencing factors for patients’ choices of rechargeable versus non-rechargeable battery

As illustrated in Figure 1, approximately half of the patients (51.6%, 99/192) who had chosen r-IPGs had a budget between 200 to 300 thousands RMB, while 53.6% (15/28) of patients with nr-IPGs only had a budget between 100 to 200 thousands RMB. The choice of r-IPG versus nr-IPG was significantly associated with the patient’s affordability (p = 0.0001). Concretely, the percentage of patients reporting ‘concern’ or ‘serious concern’ about economic issues was higher in the nr-IPG group (82.1%, 23/28) than that in the r-IPG group (60.9%, 117/192) (Figure 2D). Interestingly, the need for further surgeries to replace the battery, as well as the need for recharging the battery, was considered a concern or a serious concern by 82.1% (23/28) and by 64.3% (18/28) of the patients with nr-IPGs, respectively, also higher than r-IPG group (72.9% (140/192) and 47.9% (92/192), respectively) (Figure 2B-2C). There was 46.4% (13/28) of the patients with nr-IPGs and 51.6% (99/192) of the patients with r-IPGs reported ‘concern’ or ‘serious concern’ about the size of the battery (Figure 2A).

**Figure 1.**
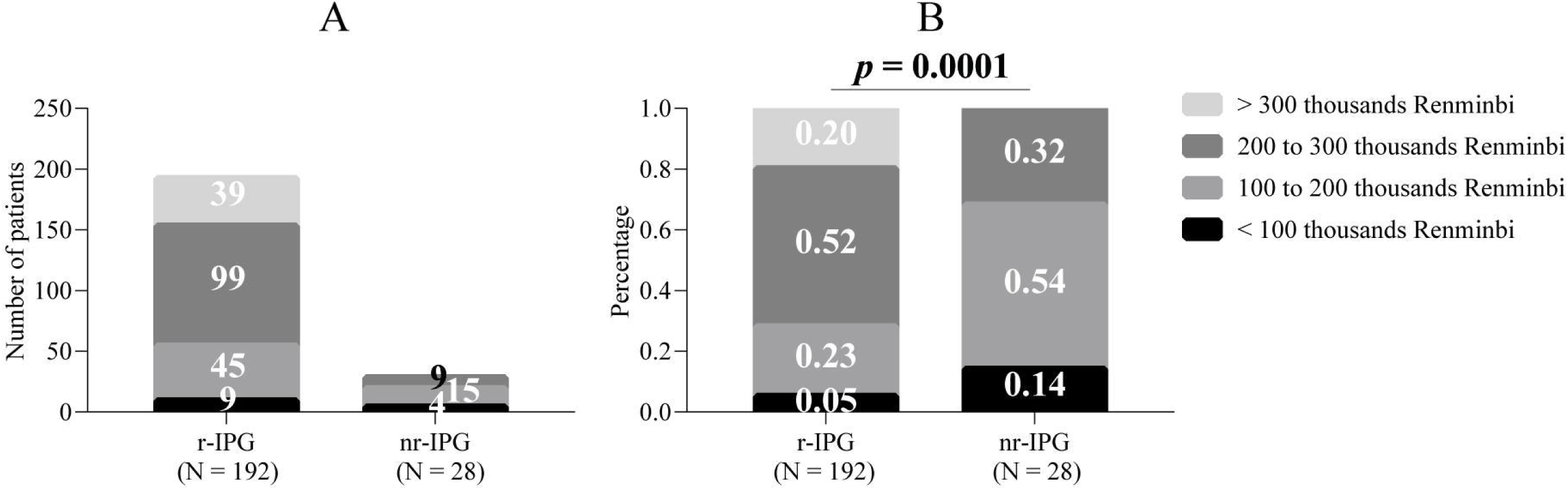
Patients’ budgets for DBS surgery. Budgets were divided into four levels and indicated by different grayscales. Values were presented either in absolute patient number (A) or in percentage (B). Abbreviation: r-IPG = rechargeable IPG (N = 192); nr-IPG: non-rechargeable IPG (N = 28).

**Figure 2.**
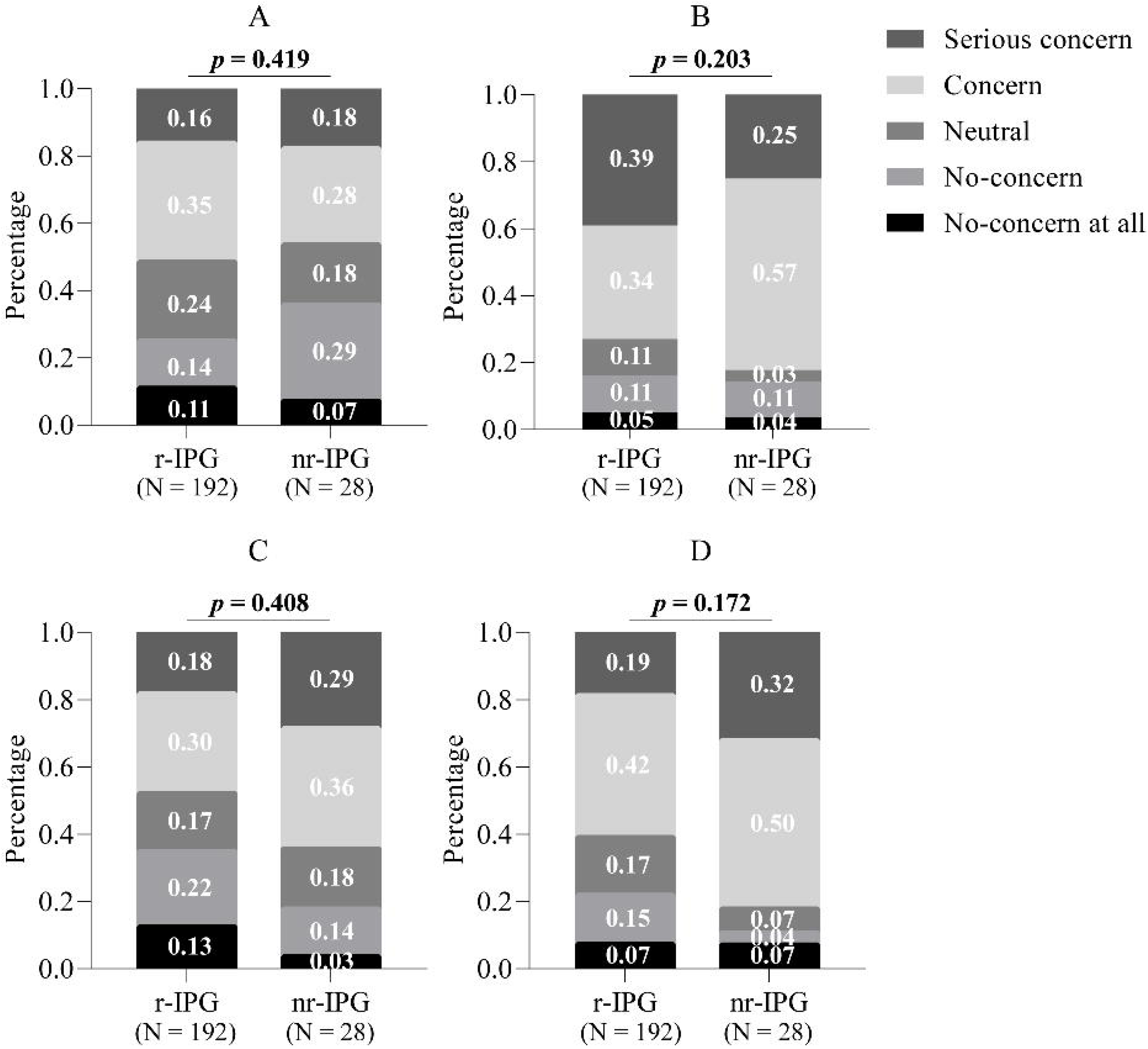
Influencing factors for choices of rechargeable versus non-rechargeable battery. The degree of influence was divided into five levels and indicated by different grayscales. Values were presented as percentage. (A) battery dimension; (B) need for further surgeries to replace the battery; (C) need for recharging the battery; (D) economic issue. Abbreviation: r-IPG = rechargeable IPG (N = 192); nr-IPG: non-rechargeable IPG (N = 28).

### Influencing factors for r-IPG patients’ choices between international and domestic manufacturers

Among 192 patients with r-IPGs, 136 (70.8%) patients had chosen a device from the international manufacturer (i.e., Medtronic), and 56 (29.2%) had chosen a device from the domestic manufacturer (i.e., PINS or SceneRay). The patient’s economic issue, the international reputation of the product, as well as the product feature of remote programming significantly affected r-IPG patients’ choices between international and domestic manufacturers (p = 6.28E-7, 5.40E-11, and 3.68E-7, respectively) (Figure 3A–3C). Not surprisingly, the patient’s choice was not influenced by clinicians (p = 0.202) (Figure 3D).

**Figure 3.**
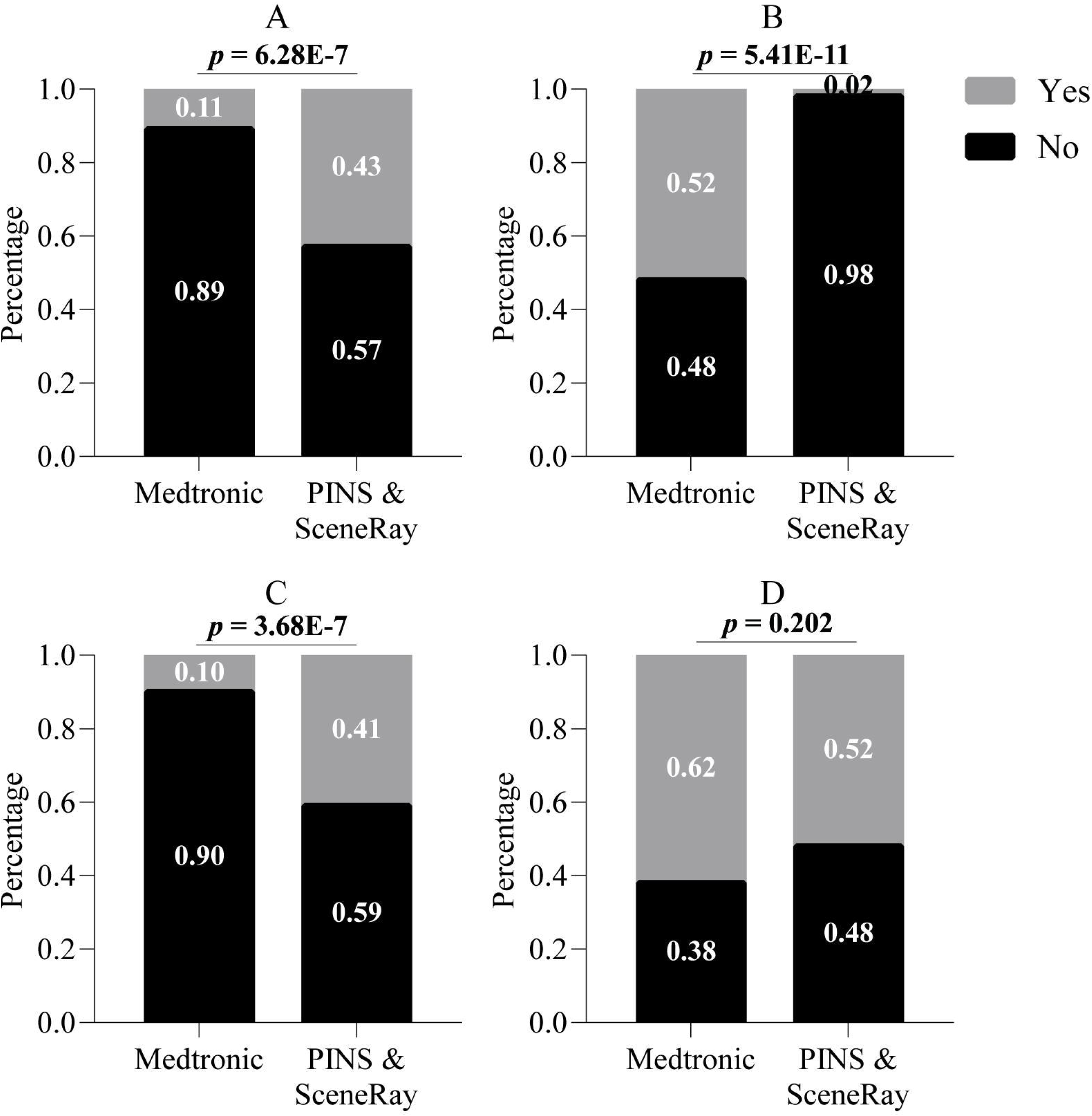
Influencing factors for choices of international versus domestic manufacturers in patients with rechargeable battery. Values were presented as percentage. (A) economic issue; (B) international reputation; (C) remote programming feature; (D) clinician’s advice. In current Chinese DBS marketing, the international manufacturer refers to Medtronic (N = 136), and domestic manufacturers refer to PINS and SceneRay (N = 56).

### Satisfaction

The majority of the patients (87.7%, 193/220) were satisfied with stimulating effects as well as the implanted device. 10% (22/220) of them claimed that their expectation had not been met after the surgery. The satisfaction rate among patients with r-IPGs (88.0% 169/192) showed no statistical significance compared with those with nr-IPGs (85.7%, 24/28) (p = 0.969) (Table 3). Also, no statistical significance was observed for the satisfaction rate between international and domestic manufacturers in patients with r-IPGs (p = 0.729). 92.7% (178/192) of the patients with r-IPGs and 92.9% (26/28) with nr-IPGs would choose the same type of device when completing the questionnaire.

**Table 3.** Satisfaction rate.

| **Questions** | **Group** |  |
| --- | --- | --- |
|  | **r-IPG** | **nr-IPG** |
|  | **(N = 192)** | **(N = 28)** |
| **1. Are you still happy with your choice of your device?** |  |  |
| Yes | 169 (88.0%) | 24 (85.7%) |
| No | 23 (12.0%) | 4 (14.3%) |
| **1.1. If not, please specify.** |  |  |
| The stimulating effects did not meet your expectation. | 18 (9.4%) | 4 (14.3%) |
| Others | 5 (2.6%) | 0 |
| **2. Would you choose the same type of your device today?** |  |  |
| Yes | 178 (92.7%) | 26 (92.9%) |
| No | 14 (7.3%) | 2 (7.1%) |

### Recharging process

The majority of the r-IPG patients (71.4%, 137/192) were capable of battery recharge independently. The remaining patients (28.6%, 55/192), however, needed partners to check and recharge the battery. Most of the patients with r-IPGs and their partners (92.7%, 178/192) felt confident of handling their device; 43.8% (78/178) of them reported the same confidence within one week after discharge from hospital. More than half of the patients (64.1%, 123/192) checked the battery every week, and most r-IPG patients (92.2%, 177/192) preferred recharging when the battery level was over 50%. The mean interval for battery recharge was 4.3 days. More than half of the patients (57.8%, 111/192) spent around 1-hour recharging. Notably, 8.3% (16/192) of the r-IPG patients reported at least one episode of recharge failure at some point, and more than half of them (68.9%, 11/16) failed to perform troubleshooting (Table 4).

**Table 4.** Recharging process for patients 1 with r-IPG. (N = 192)

| **Questions** | **Number (%)** |
| --- | --- |
| **1. Do you feel confident using your r-IPG?** |  |
| No | 14 (7.3%) |
| Yes | 178 (92.7%) |
| **1.1. If yes, how long did it take to feel confident?** |  |
| Within 1 week | 78 (40.7%) |
| 1–2 weeks | 27 (14.1%) |
| 2–4 weeks | 8 (4.2%) |
| More than 4 weeks | 65 (34.0%) |
| **2. How frequently do you check battery capacity of your r-IPG?** |  |
| Everyday | 35 (18.2%) |
| Every week | 123 (64.1%) |
| Every 2 weeks | 6 (3.1%) |
| Every 4 weeks | 17 (8.9%) |
| Every year | 11 (5.7%) |
| **3. Have you ever forgotten to recharge your r-IPG?** |  |
| No | 156 (81.3%) |
| Yes | 36 (18.7%) |
| **4. How frequently do you recharge your r-IPG?** |  |
| Everyday | 49 (25.5%) |
| 2–4 days | 28 (14.6%) |
| 5–7 days | 115 (59.9%) |
| **5. How frequently do you recharge your recharger?** |  |
| Everyday | 15 (7.8%) |
| Every week | 104 (54.2%) |
| Every 2 weeks | 31 (16.1%) |
| Every 4 weeks | 37 (19.3%) |
| Not fixed | 5 (2.6%) |
| **6. At what level of battery capacity do you usually recharge your r-IPG?** |  |
| 75–100% | 101 (52.6%) |
| 75–50% | 76 (39.6%) |
| < 50% | 15 (7.8%) |
| Warning sign | 0 |
| **7. How long does recharging usually take?** |  |
| Less than 15 min | 9 (4.7%) |
| 15–30 min | 37 (19.3%) |
| 30–45 min | 35 (18.2%) |
| 45–60 min | 40 (20.8%) |
| More than 60 min | 71 (37.0%) |
| **8. Do you check and recharge your r-IPG yourself?** |  |
| No | 55 (28.6%) |
| Yes | 137 (71.4%) |
| **9. Have you ever been unable to recharge your battery?** |  |
| No | 176 (91.7%) |
| Yes | 16 (8.3%) |
| **9.1. if yes, could you solve the problem on your own?** |  |
| No | 11 (68.9%) |
| Yes | 5 (31.2%) |

### Life with an r-IPG

During the routine recharging process, most of the r-IPG patients (92.7%, 178/192) preferred to sit or lie down to moving around. Only 26.6% (51/192) of the r-IPG patients reported at least one travel history after implantation, 78.4% (40/51) of whom recharged during their vacation. 11.5% (22/192) of the r-IPG patients continued their professional occupation after the surgery, but only one patient had recharged the IPG at work (Table 5).

**Table 5.**
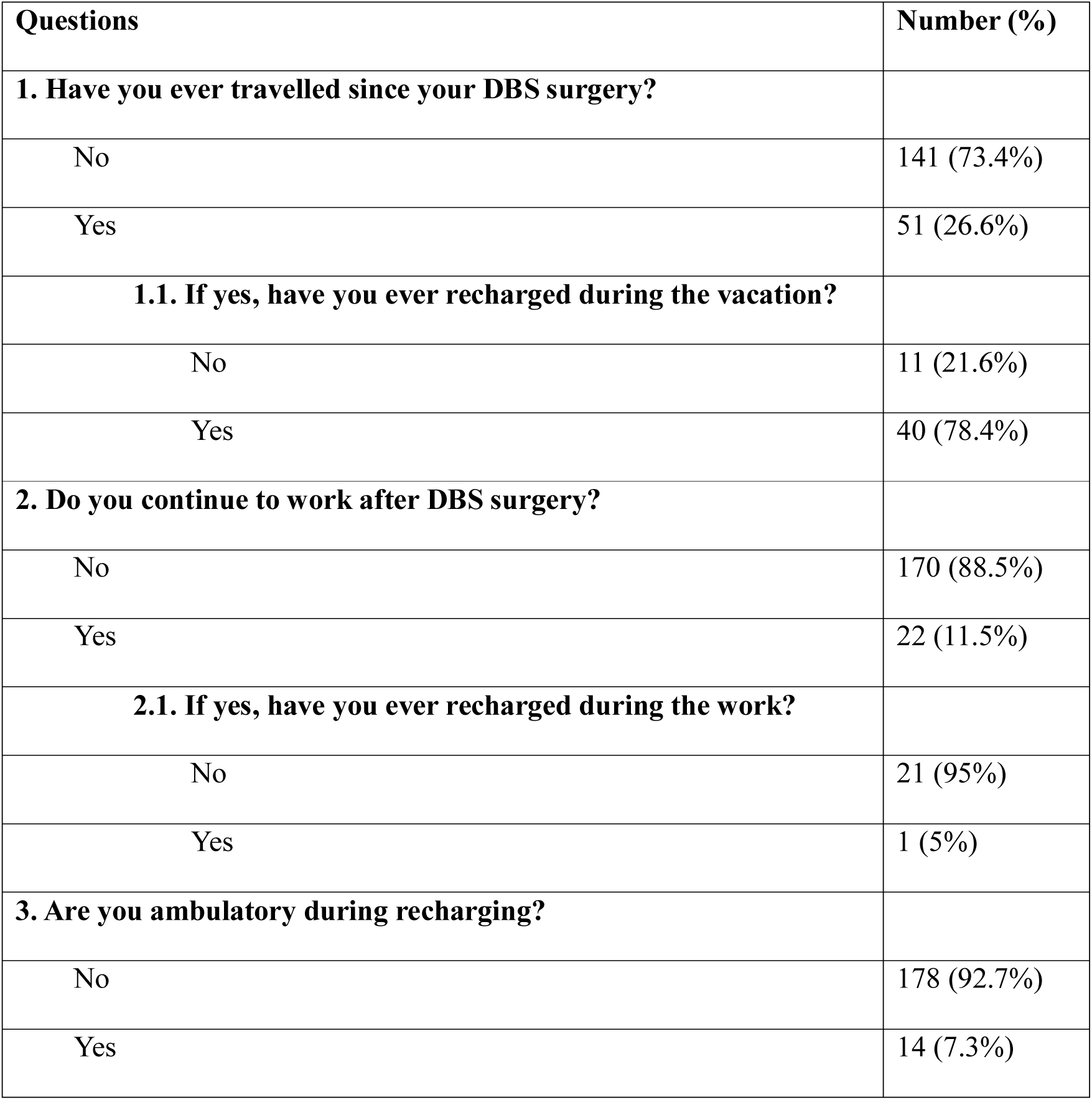
Life with a rechargeable 1 IPG. (N = 192)

## Discussion

To the best of our knowledge, our cohort is the largest one by far including all three DBS manufacturers in China and reporting the preference and satisfaction of fixed life versus rechargeable battery in PD patients. This study presented Chinese patients’ views independent of clinical influences and revealed that most patients, around 80%, opted for the rechargeable batteries. A significant factor contributing to this choice was related to economic status. Nevertheless, it is the effect of surgery other than the product being the main factor that determines patient satisfaction. The remote programming feature significantly affected patients’ choice of products, which is now only available in Chinese domestic brands. The majority of the patients with r-IPG felt confident of mastering the recharging process within a short time after discharge. We did not observe a difference regarding recharging patterns for three manufacturers. Patients mostly preferred to check and recharge the battery about once a week. The amount of time that needs to be invested by the patient to handle and recharge the IPG is around one hour. Our cohort showed that patients were able to continue their jobs and to travel with r-IPGs, but only a few of our patients recharged their IPGs during work or traveling.

While greater age, as well as more advanced functional impairment, may be considered as barriers against r-IPG utilization, neither age nor education level had a significant impact on the understanding and performance of the recharging process in the current study. Even if there exists a difficulty, patients can seek help from caregivers, as approximately 30% of r-IPG patients did in our study. Interestingly, there were several patients checked their batteries every one year or more, meaning that they recharged the IPG frequently but they did not check the IPG status as much. Although it was not reported in these participants, we had patients who failed to get timely recharge. Restarting power depleted IPGs needs continuous charging for more than 5 hours. In most cases, power depletion did not cause irreversible consequences, but one patient got aspiration pneumonia during the power-off period.

Inability to recharge the IPG could be annoying and might have serious adverse consequences. In our study, most cases’ problems were caused by malfunctioning power banks that needed to be replaced. One patient reported that the device was shut down unexpectedly without the patient’s awareness, and the program controller was used to restart the IPG. Another case involved an elderly patient, whose new caregiver failed to recharge the IPG when the original caregiver was replaced at the nursing home.

The research concerns patient’s satisfaction have been reported before. Timmermann et al.^9^ described a small prospective patient satisfaction cohort (N = 21) during a 3-month follow-up. Jakobs et al.^5^ reported another small retrospective cohort (N = 35) with a more extensive follow-up duration (mean value of 21.2 months). Their work together indicated that there was no significant association between the satisfaction rate and the number of training sessions. These two studies had limited subjects which included the patients with PD, dystonia, and essential tremor together into the analysis. Similarly, Jakobs et al. also pointed out that the majority of patients who claimed a lack of confidence after the training program had only participated in only one single session. They, therefore, proposed at least two training sessions for patients. In our study, 178 (92.7%) patients felt confident of using their IPG, and such a high rate of satisfaction is probably because our patients and their caregivers are encouraged to participate in two training sessions: one before discharge and one at one week after discharge. Waln and Jimenez-Shahed et al.^10^ reported patient satisfaction in a series of 31 patients of which 12 received r-IPGs as their initial implants. Interestingly, these patients were more satisfied with their IPGs compared with patients who previously had an nr-IPG implanted. In our study, we didn’t explore the satisfaction rates between multiple products but found the therapy efficacy was the dominating factor of patients’ general satisfaction.

Our results demonstrated that patient’s affordability had a significant impact on the choice of r-IPG versus nr-IPG in Chinese PD patients. Rizzi et al. estimated that more than 223,000 EUR could be saved over nearly 8 years in 149 patients when they were all implanted with r-IPG instead of nr-IPG^11^. However, reimbursement policies largely differ across countries and cities. As DBS electrodes and IPG are now only partly reimbursed by insurance in China, DBS therapy remains to be a substantial financial burden for many cases. The role of insurance policies, as well as other factors like the longevity of individual life expectancy, should be elucidated in the further cost-effective investigation.

We recognize some limitations in this study. First, this study was undertaken in a single institution, so there might be inherent biases. Second, as a cross-sectional study, we are not able to identify how the patient’s satisfaction developed over time as the disease progresses. Third, the follow-up duration of the current study is not sufficient to evaluate potential issues arising when r-IPGs are near the end of the recommended run-time.

In conclusion, we found most patients were satisfied with their choices of IPG. The patients’ financial status and the product feature of remote programming were the two most critical factors in their decision. The skill of using rechargeable IPG was easy to be mastered by most patients.

## Data Availability

Data included in this manuscript is all collected from patients in our center, none data from other other dataset is used in our manuscript.

## Acknowledgments

We thank Ms. Kaile Zhang and Dr. Luciano Furlanetti for their kind support in this study.

---

**Supplementary Figure 1. Size comparison between non-rechargeable (left column) and rechargeable (right column) implantable pulse generators from three manufacturers in China** (A) – (B) Medtronic; (C) – (D) PINS; (E) – (F) SceneRay.

